# COVID-19 Disease Severity among People with HIV Infection or Solid Organ Transplant in the United States: A Nationally-representative, Multicenter, Observational Cohort Study

**DOI:** 10.1101/2021.07.26.21261028

**Authors:** Jing Sun, Rena C. Patel, Qulu Zheng, Vithal Madhira, Amy L. Olex, Jessica Y. Islam, Evan French, Teresa Po-Yu Chiang, Hana Akselrod, Richard Moffitt, G. Caleb Alexander, Kathleen M. Andersen, Amanda J. Vinson, Todd T. Brown, Christopher G. Chute, Keith A. Crandall, Nora Franceschini, Roslyn B. Mannon, Gregory D. Kirk, National COVID Cohort Collaborative (N3C) Consortium

**Author notes:** Corresponding authors: Jing Sun, MD, MPH, PhD., Department of Epidemiology, Johns Hopkins University Bloomberg School of Public Health, Baltimore, MD,; Gregory D. Kirk, MD, MPH, PhD., Department of Epidemiology, Johns Hopkins University Bloomberg School of Public Health Baltimore, MD.

## Abstract

**Background:** Individuals with immune dysfunction, including people with HIV (PWH) or solid organ transplant recipients (SOT), might have worse outcomes from COVID-19. We compared odds of COVID-19 outcomes between patients with and without immune dysfunction.

**Methods:** We evaluated data from the National COVID-19 Cohort Collaborative (N3C), a multicenter retrospective cohort of electronic medical record (EMR) data from across the United States, on. 1,446,913 adult patients with laboratory-confirmed SARS-CoV-2 infection. HIV, SOT, comorbidity, and HIV markers were identified from EMR data prior to SARS-CoV-2 infection. COVID-19 disease severity within 45 days of SARS-CoV-2 infection was classified into 5 categories: asymptomatic/mild disease with outpatient care; mild disease with emergency department (ED) visit; moderate disease requiring hospitalization; severe disease requiring ventilation or extracorporeal membrane oxygenation (ECMO); and death. We used multivariable, multinomial logistic regression models to compare odds of COVID-19 outcomes between patients with and without immune dysfunction.

**Findings:** Compared to patients without immune dysfunction, PWH and SOT had a greater likelihood of having ED visits (adjusted odds ratio [aOR]: 1.28, 95% confidence interval [CI] 1.27-1.29; aOR: 2.61, CI: 2.58-2.65, respectively), requiring ventilation or ECMO (aOR: 1.43, CI: 1.43-1.43; aOR: 4.82, CI: 4.78-4.86, respectively), and death (aOR: 1.20, CI: 1.19-1.20; aOR: 3.38, CI: 3.35-3.41, respectively). Associations were independent of sociodemographic and comorbidity burden. Compared to PWH with CD4>500 cells/mm^3^, PWH with CD4<350 cells/mm^3^ were independently at 4.4-, 5.4-, and 7.6-times higher odds for hospitalization, requiring ventilation, and death, respectively. Increased COVID-19 severity was associated with higher levels of HIV viremia.

**Interpretation:** Individuals with immune dysfunction have greater risk for severe COVID-19 outcomes. More advanced HIV disease (greater immunosuppression and HIV viremia) was associated with higher odds of severe COVID-19 outcomes. Appropriate prevention and treatment strategies should be investigated to reduce the higher morbidity and mortality associated with COVID-19 among PWH and SOT.

## INTRODUCTION

Since the beginning of the COVID-19 pandemic, the United States (U.S.) has lost over half a million lives due to SARS-CoV-2 infection ^1^. People with SARS-CoV-2 infection present with a wide spectrum of disease severity, from asymptomatic infection to critical illness with high mortality ^2,3^. Identifying high risk populations during a global pandemic is key to targeting prevention messaging, monitoring disease progression, and prioritizing available vaccination and treatment.

While some patient characteristics, such as advanced age, race/ethnicity, and a high comorbidity burden, are unequivocally associated with worse COVID-19 outcomes ^2,4-8^, the relationship between immunosuppression or immunocompromised (ISC) and COVID-19 severity is less clear. Early case series and single site studies have provided conflicting results among hospitalized COVID-19 patients with HIV and suggested people with HIV (PWH) might not be at substantially elevated risk for severe outcomes ^9^. Owing to that, the initial U.S. Centers for Disease Control and Prevention (CDC) guidelines did not identify PWH as a high-risk population ^10^ and only recently modified its guidelines and prioritized vaccination for PWH ^11^. While patients with solid organ transplants (SOT) were considered to be a higher risk population ^12^, whether the higher risk is entirely driven by comorbidity burden is still under debate ^13-15^.

To date, no large-scale, nationally-representative data from the U.S. has comprehensively assessed adverse outcomes of COVID-19 by different ISC groups ^9,16^. Rapidly accessible, real-world data based on electronic medical records (EMR) from large, representative populations can provide timely and robust risk assessments, and thereby inform prioritization of critical therapies, vaccination, and targeted intervention policies. Using data on nearly 1.5 million COVID-19 cases in the National COVID-19 Cohort Collaborative (N3C), we compared risk of severe COVID-19 outcomes among PWH and SOT to patients without either condition.

## METHODS

### Design and Setting

The N3C is a centralized, harmonized, high-granularity EMR repository initiated by the National Center for Advancing Translational Science (NCATS). Design, data collection, and data harmonization methods have been described previously ^17,18^. For this analysis, data from 55 U.S. study sites contributed patient-level EMR from January 1, 2018 to May 21^st^, 2021 to the central data repository. Each study site provides demographic, medication, laboratory test, diagnoses, and vital status to the central data repository using the Observational Medical Outcomes Partnership (OMOP) as a common data model to facilitate harmonization. Data ingestion occurs daily with harmonized data released every two weeks. Deidentified data were shared through NCATS Data Enclave under a data sharing agreement.

The current study included all patients who (1) were ≥18 years old with a positive COVID-19 diagnosis between 1/1/2020 and 5/21/2021, (2) had data released for analysis in the N3C Enclave, (3) belonged to study sites with sufficient mortality and invasive ventilation data (study sites reporting over 200 patients but no hospitalizations, invasive ventilator use, or mortality cases were excluded); and (4) had data that passed initial quality check (complete data selection is reported in **Supplementary Figure 1**). The N3C Data Enclave is approved under the authority of the NIH Institutional Review Board (IRB, IRB00249128) with Johns Hopkins University School of Medicine as a central IRB for data transfer. Institutional IRB at each study site approved the study protocol or ceded to this single IRB. The current study followed the Strengthening the Reporting of Observational Studies in Epidemiology (STROBE) reporting guidelines ^19^.

### COVID-19 Case Definition

We followed the N3C COVID-19 positive definition ^17,18^ for the current study. The COVID-19 cohort definition is publicly available at GitHub ^20^. Positive cases were defined as patients with any encounter after 1/1/2020 with: 1) one of a set of *a priori*-defined SARS-CoV-2 laboratory tests, or 2) a “strong positive” diagnostic code, or 3) two “weak positive” diagnostic codes during the same encounter or on the same date. Over 99% of the COVID-19 cases in N3C were confirmed by real time PCR; <1% were confirmed by antibody testing ^17^.

### HIV, SOT, and Other Covariates

All encounters in the same health system dating back to 1/1/2018 were included to provide data on health conditions prior to COVID-19. History of HIV infection, SOT, comorbid conditions, demographics (age, sex, race and ethnicity [non-Hispanic Black, non-Hispanic White, Hispanic, others]), smoking history (never vs. ever vs. unknown), and study sites were extracted using custom concept sets developed from International Classification of Diseases (ICD) codes, Current Procedural Terminology (CPT) codes, and other medical codes (**Supplementary table S-1** lists concept sets and codes used in definitions). HIV infection was defined by (1) HIV condition (ICD codes), (2) HIV-related laboratory results (ELISA antibody test or detection of HIV viral load), (3) HIV-related medications (antiretroviral therapy, [ART]) excluding pre-exposure prophylaxis (**Supplementary Table S-1**). HIV viral load and CD4 cell counts were extracted between 1 year before and 14 days after the first COVID-19 positive results. The closest measurement of the HIV related lab values to the initial COVID-19 diagnosis was used. CD4 cell counts and HIV viral load values across different labs were harmonized with data quality evaluated by both the N3C ISC domain team and the data harmonization team. HIV viral loads were classified into: <50 (below the limit of detection), 50-1000, and >1000 copies/mL. CD4 cell counts were categorized into <350, 350-500, and >500 cells/mm^3^. SOT was defined by either procedure codes or condition codes ^21^.

Comorbid conditions (severe heart diseases [myocardial infarction or congestive heart failure], peripheral vascular disease, stroke, dementia, pulmonary diseases, rheumatic diseases, liver diseases [mild to moderate liver diseases, severe liver diseases], diabetes mellitus [diabetes or diabetes with complications], renal diseases, and cancer [cancer or metastatic cancer]) were identified prior to the initial COVID-19 diagnosis using case definitions adapted from the N3C cohort ^17^. Number of comorbidities were classified into four groups: 0, 1, 2, ≥3. Missing data on smoking history (<3%) was considered “unknown”.

### COVID-19 Outcomes

Disease severity within 45 days of the initial COVID-19 diagnosis were defined based on EMR classifying procedure and condition codes categorized into the COVID-19 Clinical Progression Scale (CPS) established by the World Health Organization (WHO) for COVID-19 clinical research ^22^. Five levels of disease severity were defined: asymptomatic or mild disease with outpatient care only (WHO severity 1-2), mild disease requiring only an emergency department (ED) visit (WHO severity 3), moderate disease with hospitalization but without invasive ventilation (WHO Severity 4-6), severe disease with hospitalization requiring invasive ventilation or extracorporeal membrane oxygenation (ECMO) treatment (WHO severity 7-9), and death (WHO severity 10). Encounters or outcomes after 45 days were not considered COVID-19-related and not included in the analysis. To account for delayed testing, we also included ED visits or hospitalizations 14 days before the initial COVID-19 diagnosis.

### Statistical Analysis

Patient characteristics at COVID-19 diagnosis were compared by ISC group status using Chi-square test for categorical variables or Wilcoxon Rank-Sum test for continuous variables. In the primary analysis, we performed multivariable multinomial logistic regression to compare the odds of each level of disease severity by ISC group (PWH only, SOT only, PWH & SOT) compared to the reference group of all others COVID-19 patients. The maximum disease severity level within 45 days of COVID-19 diagnosis was considered the final outcome in the primary analysis. Mild disease requiring only outpatient care (WHO severity 1-2) was considered the base outcome. In a separate analysis of individuals admitted to ED or hospital, we compared odds of requiring invasive ventilation by ISC groups in univariable and multivariable logistic regression models.

Among PWH, we estimated the odds of severe outcomes (hospitalization, invasive ventilation, or death) by HIV markers (i.e., categories of CD4 cell counts and HIV RNA levels). To avoid potential impact of COVID-19-related lymphopenia ^23^ on HIV markers and outcomes, we conducted sensitivity analysis using only laboratory values reported prior to COVID-19 diagnosis. Among PWH with undetectable viremia, we evaluated odds of hospitalization by their CD4 cell counts.

In all analyses, data were evaluated in three sequential models: crude estimate, model A (controlled for sociodemographic, study site, and smoking [never, ever, unknown]), and model B (controlled for covariates in model A and comorbidity burden). After ascertaining data availability, completeness, and quality, covariates incorporated into statistical models were selected based on *a priori* knowledge of relevance. All analyses were conducted in the N3C Enclave using Spark R.

## RESULTS

### Patient characteristics at COVID-19 diagnosis

Of 1,446,913 adults with a diagnosis of COVID-19 (**Table 1**), the median age was 47 years (interquartile range [IQR]: 32-61), 55.0% were female, 52.8% were non-Hispanic White, 13.7% were non-Hispanic Black, and 16.9% were current or former smokers. We identified 8,270 PWH, 11,392 SOT, and 267 with both HIV and SOT. Compared to other COVID-19 patients, PWH and SOT patients were more likely to be older, male, non-Hispanic Black, and have more comorbid conditions (**Table 1**).

**Table 1.** Patient Characteristics at COVID-19 Diagnosis.

|  | Overall | HIV- & SOT- | PWH | SOT | HIV+ & SOT+ | p-value* |
| --- | --- | --- | --- | --- | --- | --- |
| N | 1,446,913 | 1,426,984 | 8,270 | 11,392 | 267 | <0.01 |
| Age in years, median (IQR) | 47 (32-61) | 47 (32-61) | 50 (36-59) | 57 (46-66) | 58 (48-65) | <0.01 |
| Female sex, N (%) | 795,112 (55.0) | 788,127 (55.2) | 2,277 (27.5) | 4,624 (40.6) | 84 (31.5) | <0.01 |
| Race and ethnicity, N (%) |  |  |  |  |  | <0.01 |
| White non-Hispanic | 764,247 (52.8) | 755,790 (53.0) | 2,898 (35.0) | 5,475 (48.1) | 84 (31.5) |  |
| Black non-Hispanic | 197,951 (13.7) | 192,452 (13.5) | 2,732 (33.0) | 2,669 (23.4) | 98 (36.7) |  |
| Hispanic | 97,175 (6.7) | 95,683 (6.7) | 723 (8.8) | 752 (6.6) | <20 |  |
| Others | 387,540 (26.8) | 383,059 (26.8) | 1,917 (23.2) | 2,496 (21.9) | 68 (25.5) |  |
| Smoking status, N (%) |  |  |  |  |  | <0.01 |
| current or former | 245,221 (16.9) | 239,999 (16.8) | 2,101 (25.4) | 3,024 (26.5) | 97 (36.3) |  |
| non-smoker | 1,241,218 (85.8) | 1,226,299 (85.9) | 6,237 (75.4) | 8,506 (74.7) | 176 (65.9) |  |
| unknown | 39,526 (2.7) | 39,314 (2.8) | 68 (0.8) | 138 (1.2) | <20 |  |
| Number of comorbidities, N (%) |  |  |  |  |  | <0.01 |
| 0 | 1,125,998 (77.8) | 1,119,682 (78.5) | 4,261 (51.5) | 2,047 (18.0) | <20 |  |
| 1 | 177,728 (12.3) | 173,829 (12.2) | 2,066 (25.0) | 1,800 (15.8) | 33 (12.4) |  |
| 2 | 72,498 (5.0) | 68,967 (4.8) | 1,019 (12.3) | 2,454 (21.5) | 58 (21.7) |  |
| ≥3 | 70,689 (4.9) | 64,506 (4.5) | 924 (11.2) | 5,091 (44.7) | 168 (62.9) |  |
| HIV specific characteristics |  |  |  |  |  |  |
| CD4 Cell counts, cells/mm<sup>3</sup> |  |  | N = 3,574 |  | N = 86 | <0.01 |
| >500, N (%) | 2,223 (60.7) |  | 2,197 (61.5) |  | 26 (30.2) |  |
| 350-500, N (%) | 600 (16.4) |  | 587 (16.4) |  | <20 |  |
| <350, N (%) | 837 (22.9) |  | 790 (22.1) |  | 47 (54.7) |  |
| HIV viral load, copies/mL |  |  | N = 1,407 |  | N <20 | 0.27 |
| <50, N (%) | 964 (67.7) |  | 950 (67.5) |  | <20 |  |
| 50-1000, N (%) | 294 (20.6) |  | 291 (20.7) |  | <20 |  |
| >1000, N (%) | 166 (11.7) |  | 166 (11.8) |  | <20 |  |

Note: N3C policy does not allow reporting of cells with less than 20 patients, indicated as “<20”. HIV- & SOT-: people without HIV or solid organ transplant; PWH: people with HIV; SOT: solid organ transplant patient; HIV+ & SOT+: PWH had solid organ transplant; OR: odds ratio; 95% CI: 95% confidence interval.
\*P-values were based on Chi-square tests of categorical variables and Wilcoxon Rank-Sum test of continuous variables.

### COVID-19 Disease Severity among PWH and/or SOT

Within 45 days of COVID-19 diagnosis, 67.9% of patients had asymptomatic to mild disease requiring outpatient care only. While only 3.4% were solely evaluated in the ED, 26.1% required hospitalization without invasive ventilation/ECMO, 0.9% required invasive ventilation or ECMO but did not die, and 1.7% died. **Figure 1** demonstrated the crude prevalence of COVID-19 disease severity by ISC group. **Table 2** showed that patients with HIV, SOT or both had higher odds of more severe disease. Among PWH, adjusted odds ratios (ORs) for ED visits, moderate hospitalization (without invasive ventilation/ECMO), severe hospitalization with invasive ventilation/ECMO, and death were 1.28 (95% CI: 1.27, 1.29), 0.81 (95% CI: 0.78, 0.86), 1.43 (95% CI: 1.43, 1.43), and 1.20 (95% CI: 1.19, 1.20), respectively, compared to people without ISC. Compared to the crude estimates, adjustment for sociodemographic and smoking (**Table 2, model A**) and comorbidity burden (**model B**) substantially attenuated the association, but all associations remained significant. Adjustment for study sites resulted in high precision for estimations and produced narrow CI (model A and B). We observed an inverse association for moderate hospitalization (WHO severity 4-6) among PWH in adjusted models A and B, despite a high crude prevalence of PWH compared to the general population (**Figure 1**). Evaluation of individual covariates in the adjusted models indicated that the reduced odds were mainly driven by high prevalence of smoking and non-White race/ethnicity among PWH.

**Table 2.**
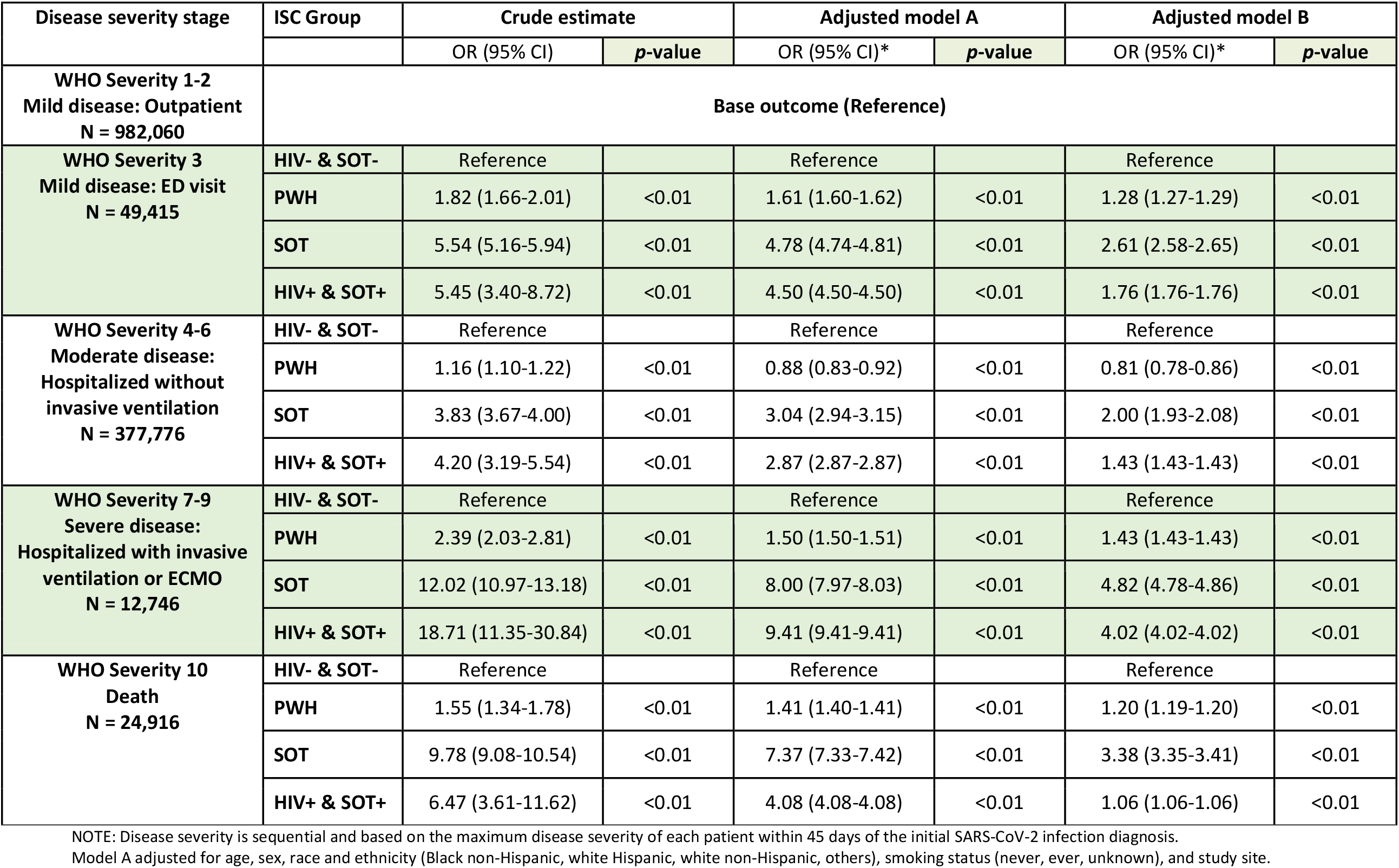
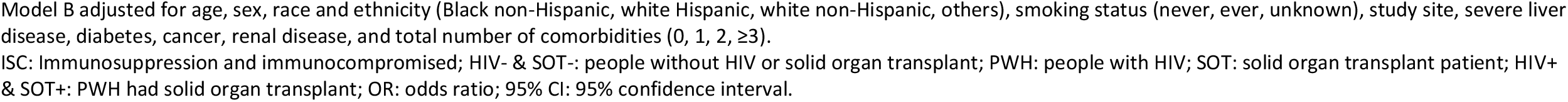
COVID-19 Disease Severity by Immunosuppression Group.

| Disease severity stage | ISC Group | Crude estimate |  | Adjusted model A |  | Adjusted model B |  |
| --- | --- | --- | --- | --- | --- | --- | --- |
|  |  | OR (95% CI) | *p*-value | OR (95% CI)* | *p*-value | OR (95% CI)* | *p*-value |
| **WHO Severity 1-2**<br>**Mild disease: Outpatient**<br>**N = 982,060** | **Base outcome (Reference)** |  |  |  |  |  |  |
| **WHO Severity 3**<br>**Mild disease: ED visit**<br>**N = 49,415** | **HIV- & SOT-** | Reference |  | Reference |  | Reference |  |
|  | **PWH** | 1.82 (1.66-2.01) | <0.01 | 1.61 (1.60-1.62) | <0.01 | 1.28 (1.27-1.29) | <0.01 |
|  | **SOT** | 5.54 (5.16-5.94) | <0.01 | 4.78 (4.74-4.81) | <0.01 | 2.61 (2.58-2.65) | <0.01 |
|  | **HIV+ & SOT+** | 5.45 (3.40-8.72) | <0.01 | 4.50 (4.50-4.50) | <0.01 | 1.76 (1.76-1.76) | <0.01 |
| **WHO Severity 4-6**<br>**Moderate disease:**<br>**Hospitalized without**<br>**invasive ventilation**<br>**N = 377,776** | **HIV- & SOT-** | Reference |  | Reference |  | Reference |  |
|  | **PWH** | 1.16 (1.10-1.22) | <0.01 | 0.88 (0.83-0.92) | <0.01 | 0.81 (0.78-0.86) | <0.01 |
|  | **SOT** | 3.83 (3.67-4.00) | <0.01 | 3.04 (2.94-3.15) | <0.01 | 2.00 (1.93-2.08) | <0.01 |
|  | **HIV+ & SOT+** | 4.20 (3.19-5.54) | <0.01 | 2.87 (2.87-2.87) | <0.01 | 1.43 (1.43-1.43) | <0.01 |
| **WHO Severity 7-9**<br>**Severe disease:**<br>**Hospitalized with invasive**<br>**ventilation or ECMO**<br>**N = 12,746** | **HIV- & SOT-** | Reference |  | Reference |  | Reference |  |
|  | **PWH** | 2.39 (2.03-2.81) | <0.01 | 1.50 (1.50-1.51) | <0.01 | 1.43 (1.43-1.43) | <0.01 |
|  | **SOT** | 12.02 (10.97-13.18) | <0.01 | 8.00 (7.97-8.03) | <0.01 | 4.82 (4.78-4.86) | <0.01 |
|  | **HIV+ & SOT+** | 18.71 (11.35-30.84) | <0.01 | 9.41 (9.41-9.41) | <0.01 | 4.02 (4.02-4.02) | <0.01 |
| **WHO Severity 10**<br>**Death**<br>**N = 24,916** | **HIV- & SOT-** | Reference |  | Reference |  | Reference |  |
|  | **PWH** | 1.55 (1.34-1.78) | <0.01 | 1.41 (1.40-1.41) | <0.01 | 1.20 (1.19-1.20) | <0.01 |
|  | **SOT** | 9.78 (9.08-10.54) | <0.01 | 7.37 (7.33-7.42) | <0.01 | 3.38 (3.35-3.41) | <0.01 |
|  | **HIV+ & SOT+** | 6.47 (3.61-11.62) | <0.01 | 4.08 (4.08-4.08) | <0.01 | 1.06 (1.06-1.06) | <0.01 |

NOTE: Disease severity is sequential and based on the maximum disease severity of each patient within 45 days of the initial SARS-CoV-2 infection diagnosis.
Model A adjusted for age, sex, race and ethnicity (Black non-Hispanic, white Hispanic, white non-Hispanic, others), smoking status (never, ever, unknown), and study site.
Model B adjusted for age, sex, race and ethnicity (Black non-Hispanic, white Hispanic, white non-Hispanic, others), smoking status (never, ever, unknown), study site, severe liver disease, diabetes, cancer, renal disease, and total number of comorbidities (0, 1, 2, $\geq 3$ ).
ISC: Immunosuppression and immunocompromised; HIV- & SOT-: people without HIV or solid organ transplant; PWH: people with HIV; SOT: solid organ transplant patient; HIV+ & SOT+: PWH had solid organ transplant; OR: odds ratio; 95% CI: 95% confidence interval.

**Figure 1.**
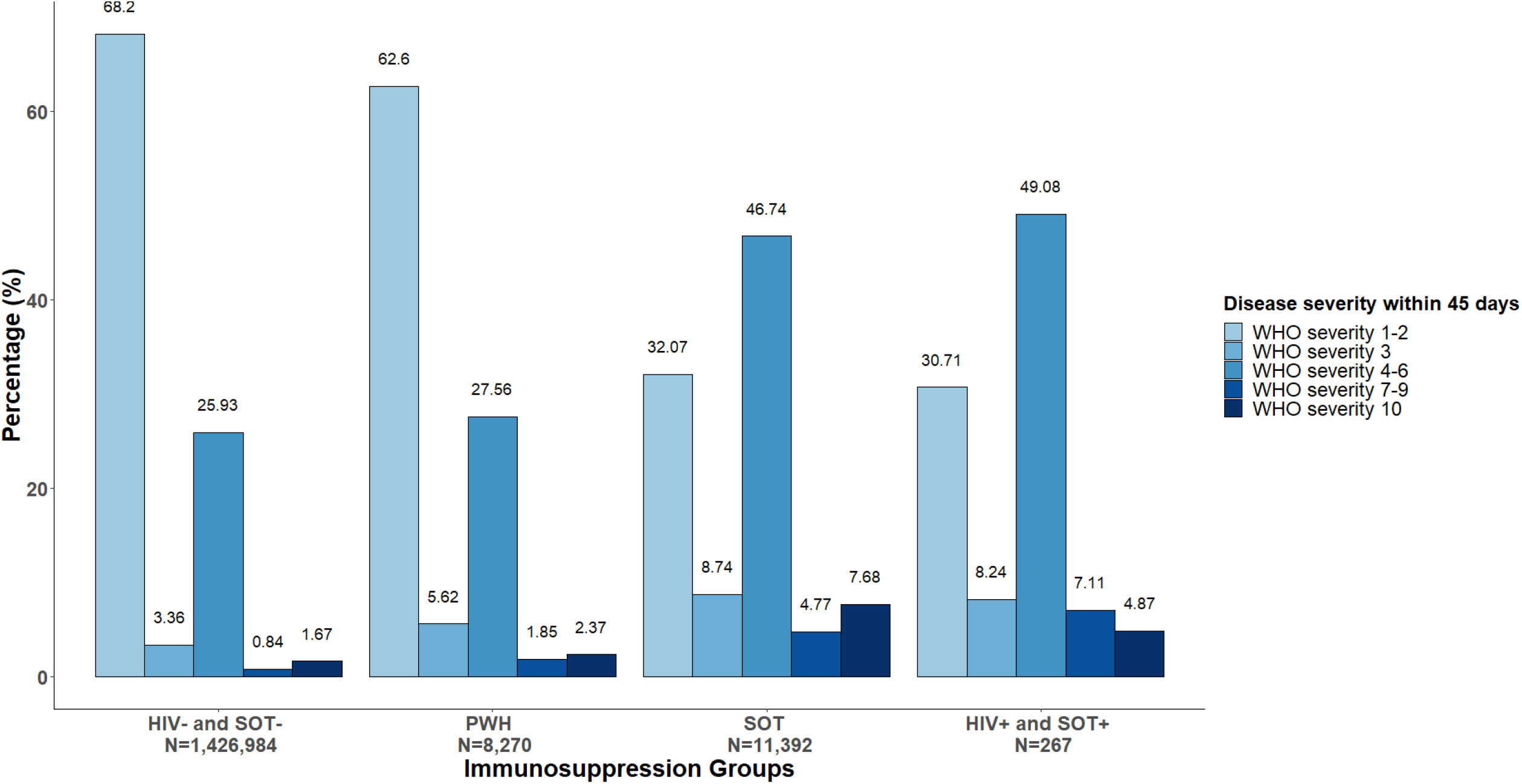
COVID-19 Disease Severity by Immunosuppression Group. HIV- & SOT-: people without HIV or solid organ transplant; PWH: people with HIV; SOT: solid organ transplant recipient; HIV+ & SOT+: PWH whom had solid organ transplant.

Among SOT patients with COVID-19, adjusted ORs for ED visit, moderate hospitalization, severe hospitalization with invasive ventilation/ECMO, and death were 2.61 (95% CI: 2.58, 2.65), 2.00 (95% CI: 1.93, 2.08), 4.82 (95% CI: 4.78, 4.86), and 3.38 (95% CI: 3.35, 3.41), respectively, compared to people without ISC. Compared to the crude estimates, associations were substantially attenuated after adjustment for sociodemographic and smoking status (**Table 2, model A**) and comorbidity burden (**model B**), but remain statistically significant. Odds of severe outcomes among PWH with SOT were similar to that observed among SOT, although the small sample size might hinder precise risk estimates.

Evaluation among COVID-19 patients admitted to ED or hospital (N = 413,292) further confirmed that the odds of requiring invasive ventilation/ECMO during their hospitalization for PWH, SOT, and people with both were 1.62 (95% CI: 1.42, 1.85), 2.29 (95% CI: 2.13, 2.47), and 2.10 (95% CI: 1.38, 3.19), respectively, compared to patients without ISC. All associations were independent of sociodemographic characteristics, smoking, and comorbidity (**Supplementary Table S-2)**.

### HIV Markers and COVID-19 Severity

A total of 3,660 PWH with COVID-19 had available CD4 cell counts measurements: 22.9% were <350 cells/mm^3^, 16.4% were between 350-500 cells/mm^3^, and 60.7% were >500 cells/mm^3^. Of 1,424 PWH with available HIV viral load measurements, the majority of them (67.7%) had undetectable viremia (<50 copies/mL), 20.6% had low level viremia (50-1000 copies/mL), and 11.7% had high level viremia (>1,000 copies/mL). Demographic characteristics of individuals with available lab values and all PWH were similar (**Supplementary Table S-3**). The associations between advanced HIV disease and COVID-19 outcomes were dose-dependent and independent of sociodemographic, smoking status, and comorbidity burdens (**Figure 2. Panel A, B, C**). Compared to CD4>500 cells/mm^3^, levels of <350 cells/mm^3^ were independently associated with greater odds of hospitalization (OR: 4.4, 95% CI: 3.6, 5.3), invasive ventilation (OR: 5.4, 95% CI: 3.2, 9), and death (OR: 7.6, 95% CI: 3.9, 14.9). Compared to undetectable viremia, viral load of 50-1000 or >1000 copies/mL were independently associated with greater odds of hospitalization (OR: 1.8 CI: 1.2-2.7 and OR: 3.5 CI: 2.2-5.5, respectively) and death (OR: 4.4 CI: 1.4-13.7 and OR: 7.3 CI: 2.1-25.7, respectively). Among PWH with undetectable viremia, lower CD4 cell counts (350-500 and <350 cells/mm ^3^) was associated with 2.9 and 6-times the odds of hospitalization (**Table 3**), respectively, compared to higher CD4 cell counts. Among PWH with CD4>500 cells/mm^3^, detectable viremia is further associated with over 2-fold higher odds of hospitalization. Sensitivity analysis restricted to laboratory values prior to COVID-19 infection yielded consistent results.

**Table 3.** Hospitalization among Well-treated People with HIV by HIV Disease Markers.

|  | Crude estimate |  | Adjusted model A |  | Adjusted model B |  |
| --- | --- | --- | --- | --- | --- | --- |
|  | OR (95% CI) | p-value | OR (95% CI) | p-value | OR (95% CI) | p-value |
| **CD4 Cell Counts among PWH with Undetectable Viremia** |  |  |  |  |  |  |
| > 500 cells/mm<sup>3</sup> | Reference |  | Reference |  | Reference |  |
| 350-500 cells/mm<sup>3</sup> | 2.7 (1.6-4.7) | <0.01 | 2.8 (1.5-5.3) | <0.01 | 2.9 (1.5-5.6) | <0.01 |
| < 350 cells/mm<sup>3</sup> | 5.3 (3.2-8.9) | <0.01 | 6.1 (3.3-11.2) | <0.01 | 6.0 (3.2-11.1) | <0.01 |
| **HIV viral load among PWH with CD4 Cell Counts > 500 cells/mm<sup>3</sup>** |  |  |  |  |  |  |
| < 50 copies/mL | Reference |  | Reference |  | Reference |  |
| ≥ 50 copies/mL | 2.3 (1.3-3.9) | <0.01 | 3.4 (1.8-6.7) | <0.01 | 3.1 (1.6-6.2) | <0.01 |

Model A adjusted for age, sex, race and ethnicity (Black non-Hispanic, white Hispanic, white non-Hispanic, others), smoking status (never, ever, unknown), and study site.
Model B adjusted for age, sex, race and ethnicity (Black non-Hispanic, white Hispanic, white non-Hispanic, others), smoking status (never, ever, unknown), study site, severe liver disease, diabetes, cancer, renal disease, and total number of comorbidities (0, 1, 2, ≥3).
Undetectable viremia: HIV viral load < 50 copies/mL.
OR: Odds Ratio; CI: Confidence Interval; PWH: People with HIV.

**Figure 2.**
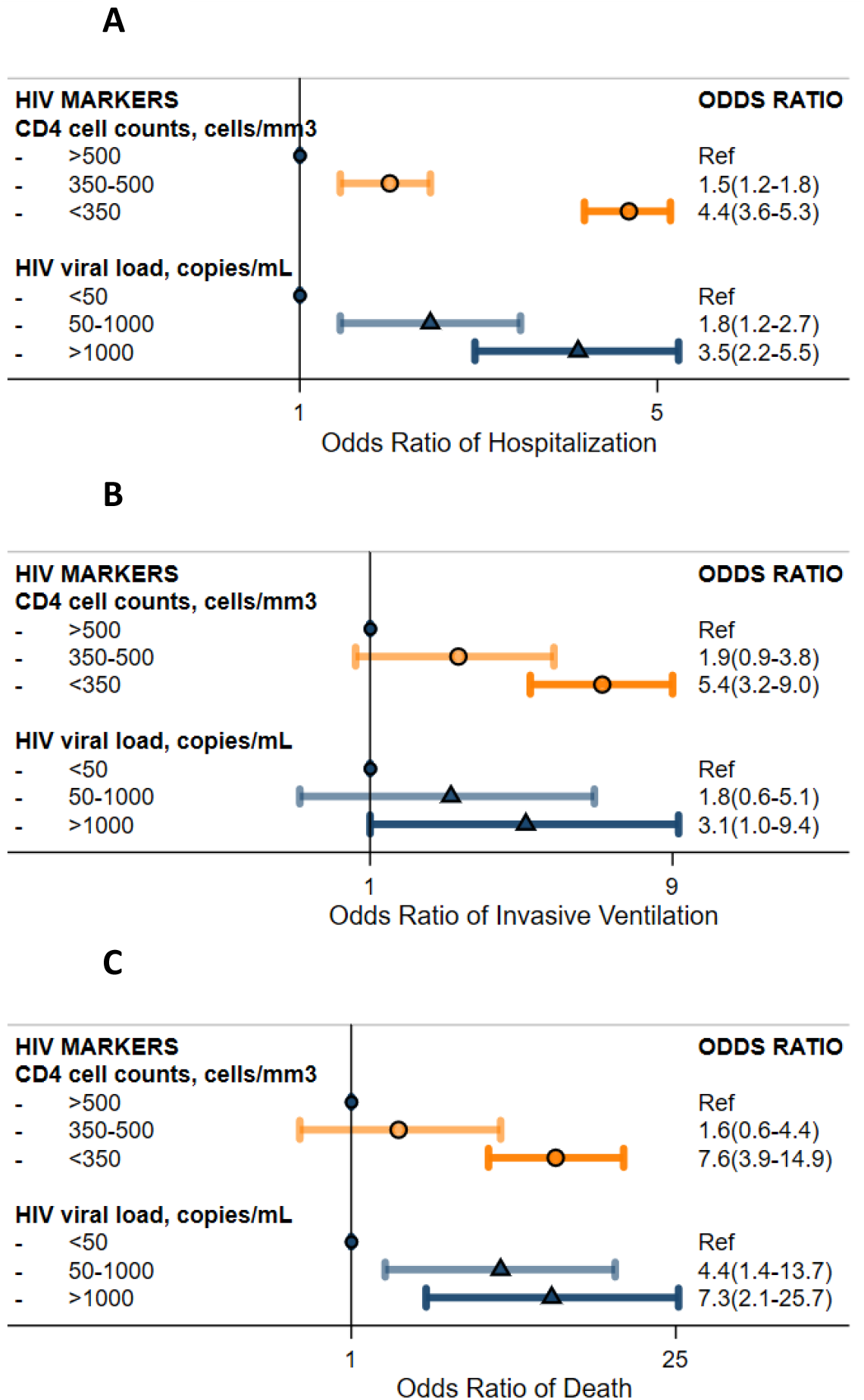
Severe COVID-19 Outcomes among People with HIV by HIV Disease Markers. Panel A. Odds of COVID-19 related hospitalization by HIV markers; Panel B. Odds of COVID-19 related invasive ventilation by HIV markers; Panel C. Odds of COVID-19 related death by HIV markers. NOTE: all outcomes (hospitalization, invasive ventilation, and death) were defined within 45 days of SARS-CoV-2 infection. Model adjusted for age, sex, race and ethnicity (Black non-Hispanic, Hispanic, white non-Hispanic, others), smoking status (never, ever, unknown), study site, severe liver disease, diabetes, cancer, renal disease, total number of comorbidities (0, 1, 2, ≥3), and study sites. *HIV markers (CD4 cell counts and HIV viral load) were extracted from EMR from 1 years before to 14 days after COVID-19 diagnosis.

## DISCUSSION

Our analysis of approximately 1.5 million U.S. patients, is among the largest and most representative multicenter cohort of COVID-19 patients assembled reflective of the U.S. pandemic to date ^17,18^. Both PWH or SOT had increased odds of severe COVID-19 outcomes, including requiring invasive ventilation/ECMO and death. The odds for severe disease were substantially driven by, though still independent of, sociodemographic variables and comorbidity burden. Among PWH, the degree of immunodeficiency and detectable HIV viremia were both independently associated with greater risk for severe COVID-19 disease. These data reveal that ISC patients are highly susceptible to severe COVID-19 and emphasize the need for identifying and implementing appropriate strategies for the prevention and management of COVID-19 disease among persons with immune dysfunction.

Whether PWH have elevated risk of severe COVID-19 disease has been under debate, and previous studies provided conflicting evidence on the association of HIV infection with COVID-19 severity ^9,15,16,24-28^. Our results add to a growing body of evidence ^13,28-31^ suggesting that PWH, indeed, have an independent increased risk for severe COVID-19 compared to the general population. One hypothesis had been that HIV populations were overrepresented with vulnerable populations to COVID-19, including increased prevalence of minorities, lower socioeconomic status, current smokers, and with substantial comorbidity. However, in sequential analysis, we continued to observe independent associations of HIV infection after adjustment for demographic variables and smoking status, as well as comorbidity burden.

While our study does not directly address the fundamental pathogenesis of how HIV may increase COVID-19 disease severity, it does provide epidemiological clues to inform the underlying biological mechanisms. Immune-dysregulation and reduced immune response to SARS-CoV-2 among PWH may increase risk of severe outcomes, given CD4 cells play a major role in antiviral immunity. Impaired regulatory T-cell response to SARS-CoV-2 appears to occur among patients with higher disease severity ^32^. Furthermore, chronic HIV-related inflammation ^33,34^ and T-cell senescence ^35^ might heighten the marked inflammatory response/cytokine storm which may intensify organ damage in severe COVID-19 disease. Indeed, our data suggested PWH with higher degree of immune dysfunction and high level of HIV viremia had a dose-dependent higher odds of severe outcomes compared to well-treated PWH. Even among PWH with undetectable viremia or with CD4>500 cells/mm ^3^, HIV viremia or immune dysfunction were both consistently associated with further risk of COVID-19 related hospitalization.

Our results call for urgent action to administer the SARS-CoV-2 vaccine to PWH to the U.S. and global population, especially targeting those individuals with poorly controlled HIV infection and those living in geographic regions where vaccine uptake among the general public has been lower ^36^. Clearly, these are not the easiest populations to reach for vaccination. Further, there has been widespread concern that the COVID-19 pandemic has negatively impacted the continuum of HIV care and prevention ^37^, even beyond the temporary disruption in HIV care visits and access to ART. The social determinants which impact an individual’s ability to successfully engage in HIV care (substance use, mental health, housing stability, comorbidity) may also be overrepresented amongst persons with poor COVID-19 outcomes. Leveraging resources from on-going HIV engagement in care initiatives and existing infrastructure (e.g., community outreach networks, “U=U”: Undetectable = Untransmittable programs) to support vaccine rollout among PWH might be critical.

Globally, these data provide the impetus to prioritize PWH for SARS-CoV-2 vaccination and consideration for early treatment where available, given the vulnerability this group faces for severe COVID-19 outcomes. Again, existing HIV care and prevention infrastructure (PEPFAR, Global Fund) will need to be leveraged to effectively reach this target population.

Our study confirmed the strong associations between SOT and severe COVID-19 reported in previous studies ^13-15^. Importantly, our data showed that such associations were independent of comorbid conditions and advanced age. It is still unclear whether immunosuppressive drugs are the driver of the elevated risk of severe COVID-19 outcomes among SOT. A previous single-center study suggested immunosuppressant use among SOT neither increased nor decreased risk of severe COVID-19 ^38^. Detailed SOT factors (e.g., type of transplant, time since transplant, specific immunosuppressive regimens) and their impact on COVID-19 is the subject of ongoing analyses ^21^ in the N3C ISC domain team. Regardless, health care providers may need to consider earlier or more permissive use of available therapies in SOT during acute COVID-19 disease ^39^.

Recent studies indicated that SOT patients experienced weak immune responses to COVID-19 vaccination ^40,41^, while three-doses of mRNA vaccine may improve the immunogenicity of the vaccine ^42,43^. Although the precise immune response among PWH after vaccination requires further evaluation, booster doses of vaccine should be considered among both SOT and PWH, especially those with advanced immunosuppression. In addition to vaccination strategies, many other management issues surrounding SARS-CoV-2 exposure and infection among ISC patients requires further detailed investigation. While most of the U.S. population is acclimating to life without COVID-19 mitigation measures in place, ISC patients and their caregivers should consider the appropriateness of continuing non-pharmaceutical interventions, including mask wearing, social distancing in personal, work, and clinical settings, avoiding travel, etc. Among ISC patients infected with or exposed to SARS-CoV-2, post-exposure prophylaxis / pre-emptive therapy, close monitoring for early disease progression, permissive use of additional therapies, evaluation of the duration of viral shedding and potential for onward transmission, and follow-up for post-acute sequelae of COVID-19 should be considered.

Despite the largest and the most representative sample to date for a U.S. population ^17,18^, our study has limitations. First, N3C primarily compiled data from large academic medical centers funded by NCATS. Therefore, it may be less representative of admission practices and disease severity in other settings. There has historically been a low threshold to admit PWH and SOT to these hospital because of their underlying disease. While this might have contributed somewhat to increased likelihood of hospitalization with mild/moderate disease, it would not account for the independent increased odds for ventilation among persons that were hospitalized in these groups. Potential misclassification of key exposures (e.g. HIV status, SOT, or other comorbidities) may occur due to inherent limitations of EMR data. In particular, we were more likely to identify PWH who were successfully engaged in care due to reliance on ICD codes, ART, and laboratory results in the past 2 years to define HIV status. However, HIV or SOT are unlikely to be missed on admission diagnoses due to their clinical importance. Further, these potential misclassifications are likely to be non-differential throughout the cohort and unlikely to change our conclusions. HIV laboratory values (CD4 cell counts and HIV viral load) were not universally available across all study sites and individuals. Differential laboratory assays and protocols across sites added further complexity for data harmonization and validation. However, further evaluation suggested that demographic characteristics of individuals with available lab values and all PWH were similar. Notably, the prevalence of HIV viral suppression among PWH in N3C was similar to U.S. CDC surveillance data (67.7% vs. 62.7%). ^44^ ISC patients may receive prioritized treatment compared to the general population, which if therapies were beneficial, could result in our study underestimating the effect size of ISC conditions and severe COVID-19. Lastly, we only evaluated ISC patients with SOT and PWH and classified patients with other immune dysfunction (e.g. multiple sclerosis, rheumatoid arthritis) as patients without ISC. Thus, we might overestimate the risk of severe COVID-19 outcomes among patients without ISC, and the risk of severe outcomes among SOT and PWH might be even higher.

Notwithstanding these limitations, our study has notable strengths. To our knowledge, this analysis included patient-level data on amongst the largest sample size of patients with ISC reported to date, which afforded sufficient statistical power to perform analyses that individual study sites are unable to. Additionally, using a harmonized multicenter data source reduces potential selection bias seen in single-center or regional studies.

In conclusion, PWH and SOT patients were highly vulnerable to SARS-CoV-2 infection and developed more severe COVID-19 outcomes compared to the general population. Immune dysfunction was associated with elevated risk of severe outcomes independent of age, smoking, and comorbidity burden. Among PWH, risk was further driven by advanced HIV disease. These results call for urgent action for targeted vaccine rollout and potential booster vaccinations among ISC patients. They also call for early treatment, close monitoring for disease progression, as well as pharmaceutical and non-pharmaceutical interventions to reduce COVID-19 risk among ISC patients during the on-going global COVID-19 pandemic.

## Data Availability

The N3C is an open data repository. All data presented in this manuscript has been approved by N3C and is publically available.

## Acknowledgements

### N3C Attribution

The analyses described in this publication were conducted with data or tools accessed through the NCATS N3C Data Enclave covid.cd2h.org/enclave and supported by NCATS U24 TR002306. This research was possible because of the patients whose information is included within the data from participating organizations (covid.cd2h.org/dtas) and the organizations and scientists (covid.cd2h.org/duas) who have contributed to the on-going development of this community resource (cite this https://doi.org/10.1093/jamia/ocaa196).

### IRB

The N3C data transfer to NCATS is performed under a Johns Hopkins University Reliance Protocol # IRB00249128 or individual site agreements with NIH. The N3C Data Enclave is managed under the authority of the NIH; information can be found at https://ncats.nih.gov/n3c/resources.

### Individual acknowledgements for Core Contributors

We gratefully acknowledge contributions from the following N3C core teams

- Principal Investigators: Melissa A. Haendel*, Christopher G. Chute*, Kenneth R. Gersing, Anita Walden
- Workstream, subgroup and administrative leaders: Melissa A. Haendel*, Tellen D. Bennett, Christopher G. Chute, David A. Eichmann, Justin Guinney, Warren A. Kibbe, Hongfang Liu, Philip R.O. Payne, Emily R. Pfaff, Peter N. Robinson, Joel H. Saltz, Heidi Spratt, Justin Starren, Christine Suver, Adam B. Wilcox, Andrew E. Williams, Chunlei Wu
- Key liaisons at data partner sites
- Regulatory staff at data partner sites
- Individuals at the sites who are responsible for creating the datasets and submitting data to N3C
- Data Ingest and Harmonization Team: Christopher G. Chute*, Emily R. Pfaff*, Davera Gabriel, Stephanie S. Hong, Kristin Kostka, Harold P. Lehmann, Richard A. Moffitt, Michele Morris, Matvey B. Palchuk, Xiaohan Tanner Zhang, Richard L. Zhu
- Phenotype Team (Individuals who create the scripts that the sites use to submit their data, based on the COVID and Long COVID definitions): Emily R. Pfaff*, Benjamin Amor, Mark M. Bissell, Marshall Clark, Andrew T. Girvin, Stephanie S. Hong, Kristin Kostka, Adam M. Lee, Robert T. Miller, Michele Morris, Matvey B. Palchuk, Kellie M. Walters
- Project Management and Operations Team: Anita Walden*, Yooree Chae, Connor Cook, Alexandra Dest, Racquel R. Dietz, Thomas Dillon, Patricia A. Francis, Rafael Fuentes, Alexis Graves, Julie A. McMurry, Andrew J. Neumann, Shawn T. O’Neil, Usman Sheikh, Andréa M. Volz, Elizabeth Zampino
- Partners from NIH and other federal agencies: Christopher P. Austin*, Kenneth R. Gersing*, Samuel Bozzette, Mariam Deacy, Nicole Garbarini, Michael G. Kurilla, Sam G. Michael, Joni L. Rutter, Meredith Temple-O’Connor
- Analytics Team (Individuals who build the Enclave infrastructure, help create codesets, variables, and help Domain Teams and project teams with their datasets): Benjamin Amor*, Mark M. Bissell, Katie Rebecca Bradwell, Andrew T. Girvin, Amin Manna, Nabeel Qureshi
- Publication Committee Management Team: Mary Morrison Saltz*, Christine Suver*, Christopher G. Chute, Melissa A. Haendel, Julie A. McMurry, Andréa M. Volz, Anita Walden
- Publication Committee Review Team: Carolyn Bramante, Jeremy Richard Harper, Wenndy Hernandez, Farrukh M Koraishy, Federico Mariona, Saidulu Mattapally, Amit Saha, Satyanarayana Vedula

### Additional data partners who have signed DTA and data release pending

The Rockefeller University — UL1TR001866: Center for Clinical and Translational Science • The Scripps Research Institute — UL1TR002550: Scripps Research Translational Institute • University of Texas Health Science Center at San Antonio — UL1TR002645: Institute for Integration of Medicine and Science • The University of Texas Health Science Center at Houston — UL1TR003167: Center for Clinical and Translational Sciences (CCTS) • NorthShore University HealthSystem — UL1TR002389: The Institute for Translational Medicine (ITM) • Yale New Haven Hospital — UL1TR001863: Yale Center for Clinical Investigation • Emory University — UL1TR002378: Georgia Clinical and Translational Science Alliance • Weill Medical College of Cornell University — UL1TR002384: Weill Cornell Medicine Clinical and Translational Science Center • Montefiore Medical Center — UL1TR002556: Institute for Clinical and Translational Research at Einstein and Montefiore • Medical College of Wisconsin — UL1TR001436: Clinical and Translational Science Institute of Southeast Wisconsin • University of New Mexico Health Sciences Center — UL1TR001449: University of New Mexico Clinical and Translational Science Center • George Washington University — UL1TR001876: Clinical and Translational Science Institute at Children’s National (CTSA-CN) • Stanford University — UL1TR003142: Spectrum: The Stanford Center for Clinical and Translational Research and Education • Regenstrief Institute — UL1TR002529: Indiana Clinical and Translational Science Institute • Cincinnati Children’s Hospital Medical Center — UL1TR001425: Center for Clinical and Translational Science and Training • Boston University Medical Campus — UL1TR001430: Boston University Clinical and Translational Science Institute • The State University of New York at Buffalo — UL1TR001412: Clinical and Translational Science Institute • Aurora Health Care — UL1TR002373: Wisconsin Network For Health Research • Brown University — U54GM115677: Advance Clinical Translational Research (Advance-CTR) • Rutgers, The State University of New Jersey — UL1TR003017: New Jersey Alliance for Clinical and Translational Science • Loyola University Chicago — UL1TR002389: The Institute for Translational Medicine (ITM) • #N/A — UL1TR001445: Langone Health’s Clinical and Translational Science Institute • Children’s Hospital of Philadelphia — UL1TR001878: Institute for Translational Medicine and Therapeutics University of Kansas Medical Center — UL1TR002366: Frontiers: University of Kansas Clinical and Translational Science Institute • Massachusetts General Brigham — UL1TR002541: Harvard Catalyst • Icahn School of Medicine at Mount Sinai — UL1TR001433: ConduITS Institute for Translational Sciences • Ochsner Medical Center — U54GM104940: Louisiana Clinical and Translational Science (LA CaTS) Center • HonorHealth — None (Voluntary) • University of California, Irvine — UL1TR001414: The UC Irvine Institute for Clinical and Translational Science (ICTS) • University of California, San Diego — UL1TR001442: Altman Clinical and Translational Research Institute • University of California, Davis — UL1TR001860: UCDavis Health Clinical and Translational Science Center • University of California, San Francisco — UL1TR001872: UCSF Clinical and Translational Science Institute • University of California, Los Angeles — UL1TR001881: UCLA Clinical Translational Science Institute • University of Vermont — U54GM115516: Northern New England Clinical & Translational Research (NNE-CTR) Network • Arkansas Children’s Hospital — UL1TR003107: UAMS Translational Research Institute

## Supplementary Materials

**Supplementary Figure 1.**
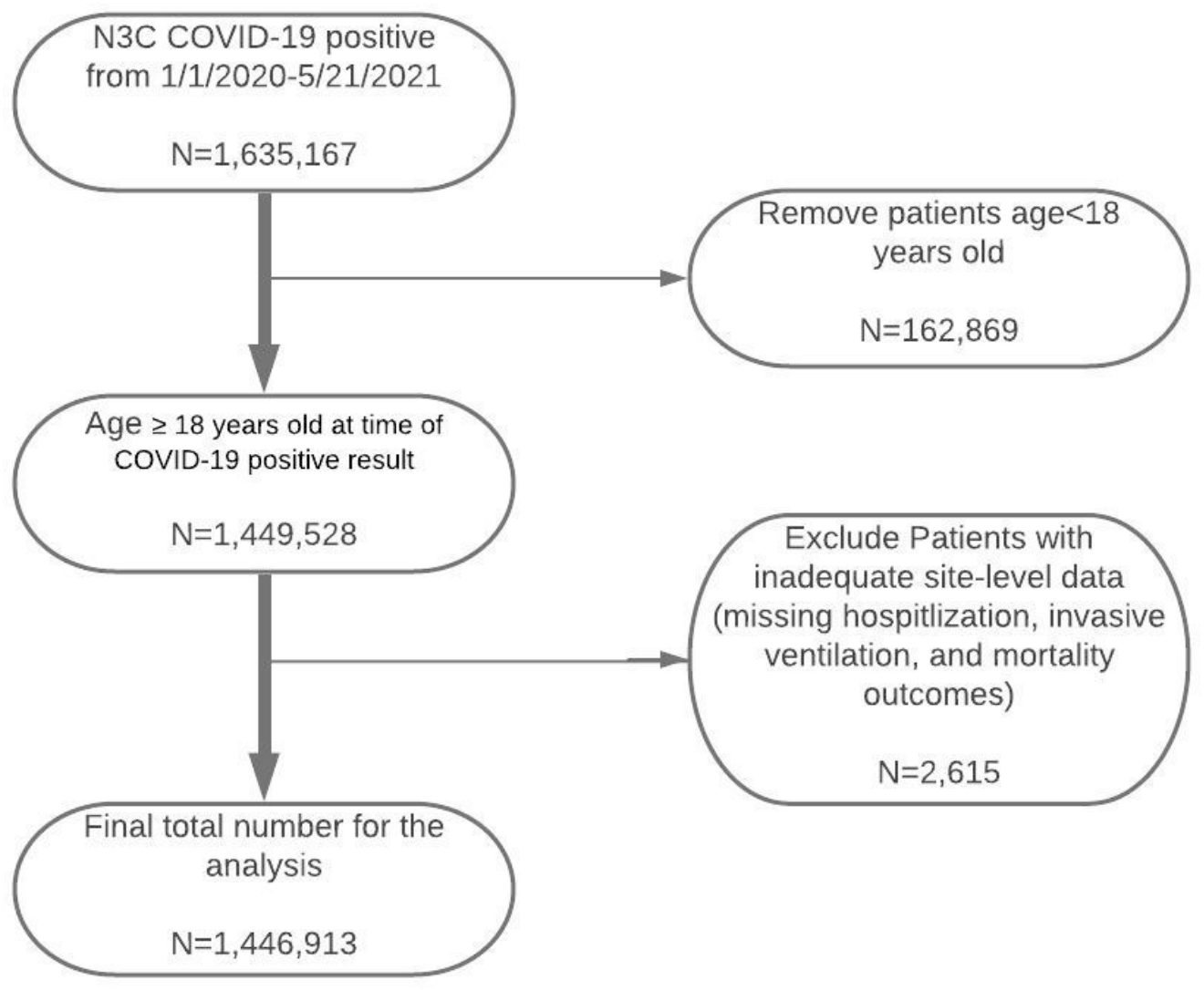
Flow Diagram of Study Sample.

**Supplementary Table S-1.**
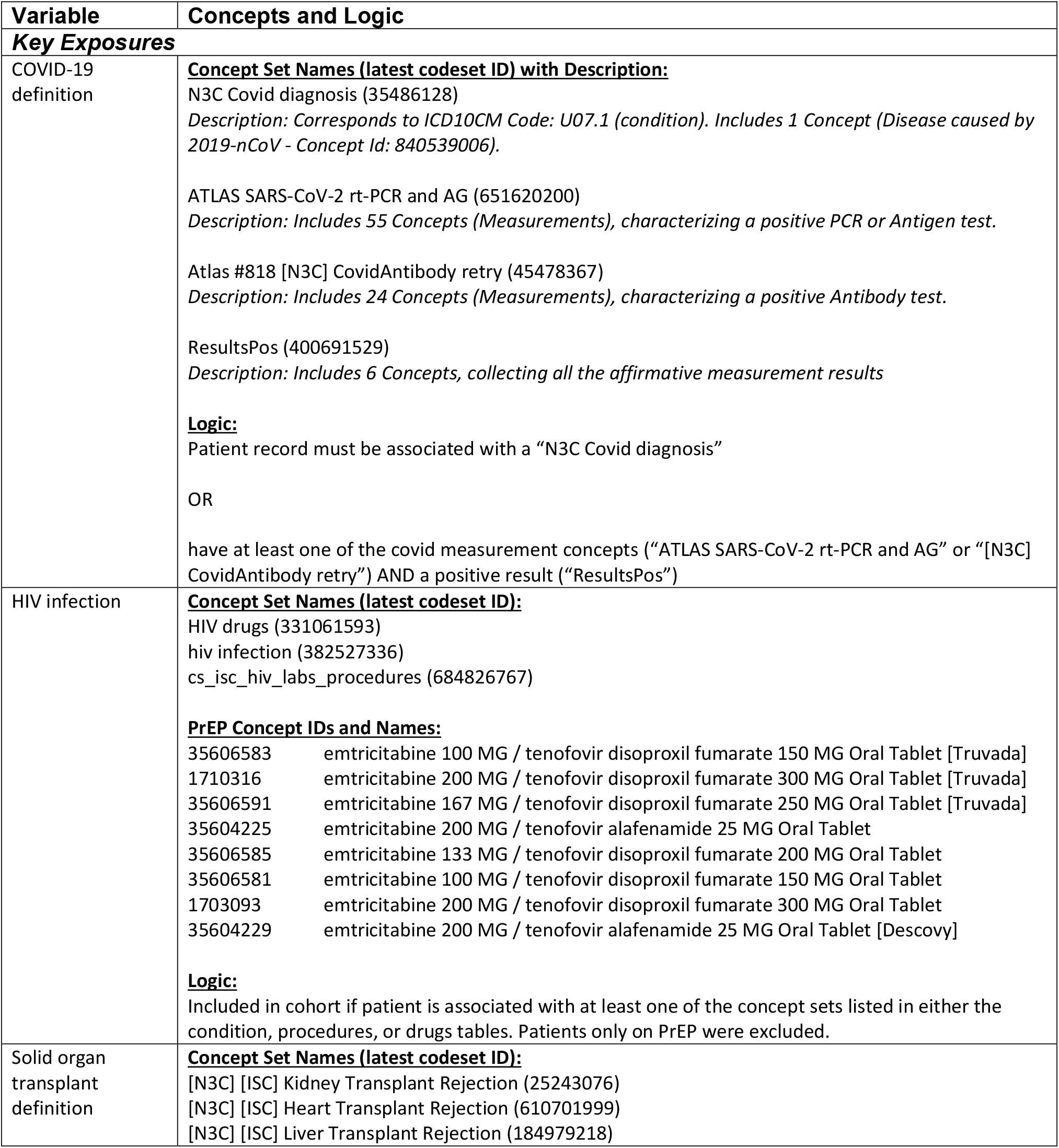
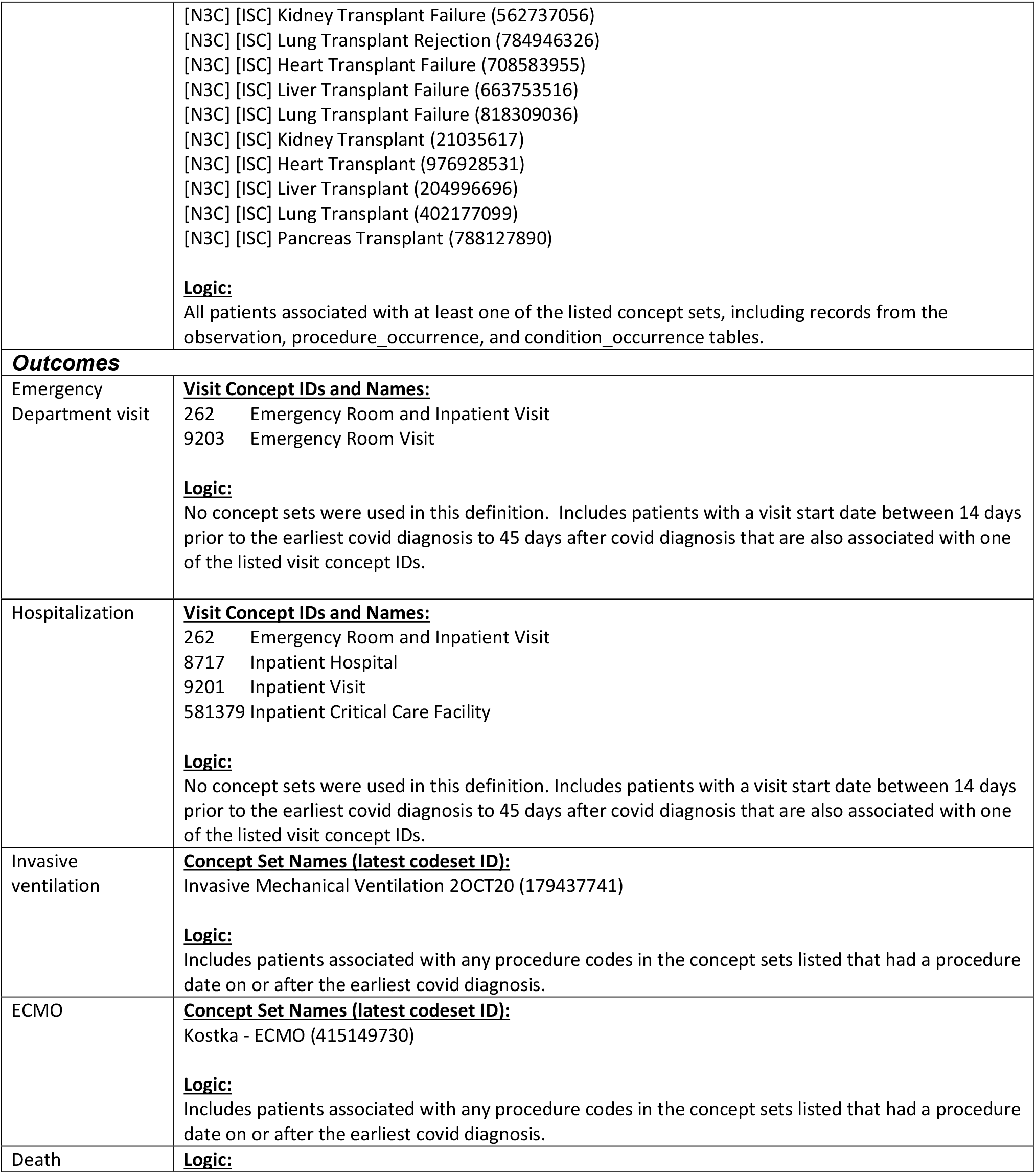
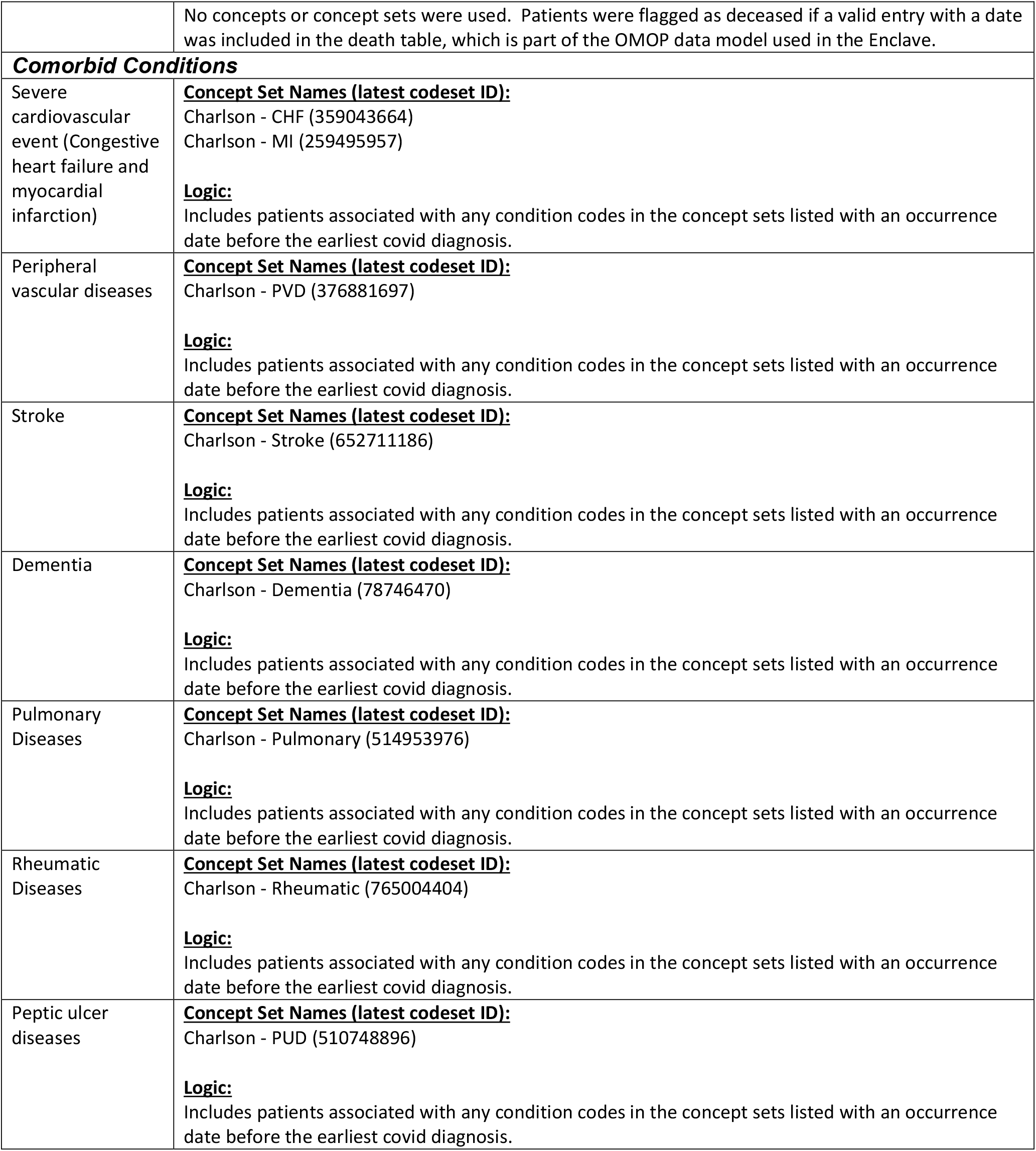
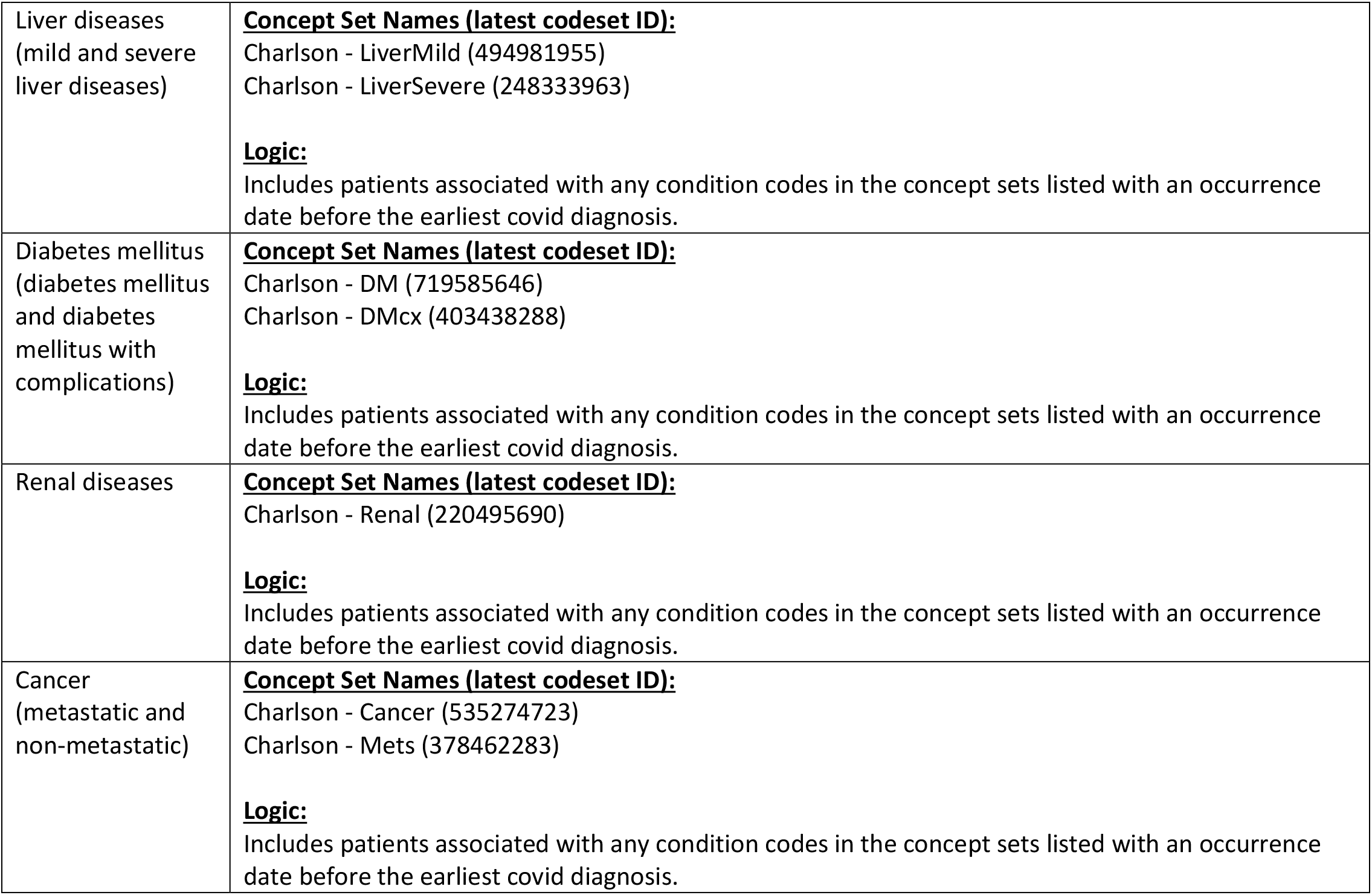
Key Variables and Concept Definitions.

**Supplementary Table S-2.** Invasive Mechanical Ventilation Use among COVID-19 Patients Admitted to Emergency Department or Hospital by Immunosuppression group.

| Immunosuppression group<br>N = 413,292 | Crude estimates |  | Adjusted estimates A |  | Adjusted estimates B |  |
| --- | --- | --- | --- | --- | --- | --- |
|  | OR (95% CI) | P-value | OR (95% CI) | P-value | OR (95% CI) | P-value |
| **HIV- & SOT- (N = 403,847)** | Ref. | Ref. | Ref. | Ref. | Ref. | Ref. |
| **PWH (N = 2,611)** | 1.85 (1.62-2.11) | <0.01 | 1.66 (1.46-1.90) | <0.01 | 1.62 (1.42-1.85) | <0.01 |
| **SOT (N = 6,671)** | 3.26 (3.05-3.49) | <0.01 | 2.86 (2.67-3.07) | <0.01 | 2.29 (2.13-2.47) | <0.01 |
| **HIV+ & SOT+ (N = 163)** | 3.63 (2.41-5.45) | <0.01 | 2.88 (1.9-4.36) | <0.01 | 2.10 (1.38-3.19) | <0.01 |

Model A adjusted for age, sex, race and ethnicity (Black non-Hispanic, white Hispanic, white non-Hispanic, others), smoking status, and study site.
Model B adjusted for age, sex, race and ethnicity (Black non-Hispanic, white Hispanic, white non-Hispanic, others), smoking status, study site, severe liver disease, diabetes, cancer, renal disease, and number of comorbidities (0, 1, 2, ≥3).
HIV- & SOT-: people without HIV or solid organ transplant; PWH: people with HIV; SOT: solid organ transplant patient; HIV+ & SOT+: PWH had solid organ transplant; OR: odds ratio; 95% CI: 95% confidence interval.

**Supplementary Table S-3.** Characteristics of People with HIV by Their Laboratory Data Availability.

|  | **PWH with**<br>**CD4 cell**<br>**count data** | **PWH with**<br>**HIV viral**<br>**load data** | **all PWH\*** |
| --- | --- | --- | --- |
| **N** | N = 3,660 | N = 1,424 | N = 8,537 |
| **age, median (IQR)** | 52 (39-60) | 52 (39-60) | 50 (37-59) |
| **Age ≥ 60, N (%)** | 946 (25.8) | 385 (27) | 2,087 (24.4) |
| **female sex, N (%)** | 871 (23.8) | 307 (21.6) | 2361 (27.7) |
| **race and ethnicity, N (%)** |  |  |  |
| **White non-Hispanic** | 1117 (30.5) | 1,117 (30.5) | 2982 (34.9) |
| **Black non-Hispanic** | 1200 (32.8) | 405 (28.4) | 2830 (33.1) |
| **Hispanic** | 421 (11.5) | 229 (16.1) | 681 (9.4) |
| **Others** | 922 (25.2) | 274 (19.2) | 1,985 (23.3) |
| **smoking status, N (%)** |  |  |  |
| **current or former** | 1985 (23.3) | 430 (30.2) | 2198 (25.7) |
| **non-smoker** | 2,198 (25.7) | 994 (69.8) | 6413 (75.1) |
| **unknown** | 35 (1) | <20 | 74 (0.9) |
| **number of comorbidities, N (%)** |  |  |  |
| **0** | 1572 (43) | 576 (40.4) | 4,269 (50) |
| **1** | 1,047 (28.6) | 442 (31) | 2,099 (24.6) |
| **2** | 533 (14.6) | 225 (15.8) | 1,077 (12.6) |
| **≥ 3** | 508 (13.9) | 181 (12.7) | 1,092 (12.8) |

Note: N3C policy does not allow reporting of cells with less than 20 patients, indicated as "<20".
PWH=people with HIV
\*All PWH included people with HIV alone and people with HIV and solid organ transplant.

